# Cellular and Antibody Immunity after COVID-19 Vaccination at >4-Month Follow Up in Immunocompetent and Immunocompromised Subjects

**DOI:** 10.1101/2021.10.07.21257459

**Authors:** Rakesh Sindhi, Chethan Ashokkumar, Vineeta Singh, Brianna Spishock, Maggie Saunders, Angelo Mabasa, Elizabeth Sindhi, Pradeep Sethi, Ashok Reddy, Shankar Subramaniam, Bobby Nibhanupudy

## Abstract

We evaluated post-vaccination immunity after COVID-19 vaccination with serial changes in cellular and antibody responses to the spike protein S, its S2 component which is conserved between SARS-CoV-2 and human coronaviruses, and the S1 component, which is specific to SARS-CoV-2 and also contains its receptor binding domain (RBD). In 21 healthy immunocompetent subjects all of whom demonstrated circulating IgG antibodies 4 months after mRNA1273 or BNT162b vaccination, a) the strength of S-IgG was stable while RBD-IgG declined, b) S2-reactive B-cell frequencies increased progressively (p=0.002) c) S1-reactive CD8+T-cells and CD19+B-cells were undetectable after a transient increase, and d) monocytic and polymorphonuclear myeloid-derived suppressor cells (M-MDSC, PMN-MDSC) increased after the first vaccine dose. Compared with 4-month measurements from immunocompetent subjects, single samples from 20 vaccinated immunocompromised (IC) subjects revealed a) circulating S-IgG and RBD-IgG in 13 (65%) and 9 (45%) subjects, respectively, b) no differences in S2-reactive T- and B-cells, c) undetectable S1-reactive T- and B-cells, and d) fewer S-reactive CD8+T-cells and CD19+B-cells (p<0.05). Among 11 IC recipients who failed to make RBD-IgG, frequencies of PMN-MDSC were significantly higher (p<0.0004) compared with IC or immunocompetent subjects with RBD-IgG. COVID-19 vaccination induces stable antibodies to the spike protein and expands circulating B-cells reactive to the conserved spike protein sequence in immunocompetent subjects. MDSC which are known to suppress T- and B-cells, and which increase after vaccination, may limit post-vaccination responses especially among immunocompromised subjects. Antibody and cellular responses to SARS-CoV-2-specific spike antigenic sequences appear to be less durable.

## INTRODUCTION

Cellular immunity to the COVID-19 coronavirus is of great interest because of concerns about vaccine efficacy against emerging SARS-CoV-2 variants and the lower seroconversion rates after vaccination in older and immunosuppressed subjects (1-3). Following COVID-19 vaccination, antibodies only develop in a half of transplant recipients, inflammatory bowel disease patients receiving anti-tumor necrosis factor antibodies, and those with hematologic malignancies (3-5). The seroconversion rate is only 23% in those with B-cell malignancies (5).

The post-vaccination immune response likely recapitulates the discordant kinetics of T-cell responsiveness and SARS-CoV-2 antibodies observed by us early after moderate to severe COVID-19 infection (6). Although anti-spike IgG antibodies appeared within two weeks after COVID-19 infection in immunocompetent subjects, T-cells from severely ill COVID-19 patients displayed impaired in vitro reactivity to the spike antigen, while circulating myeloid derived suppressor cells (MDSC) increased significantly. MDSC arise during acute viral infections and suppress T-cells (7-9). In immunosuppressed transplant recipients, we observed a lower incidence of antibody to the SARS-CoV-2-specific receptor binding domain (RBD) of the spike protein, significantly impaired T-cell responses to stimulation with conserved spike antigen, and a 34% incidence of CMV co-infection early after COVID-19 infection. The S antigenic sequence consists of the less conserved N-terminal S1 component, which contains the receptor binding domain (RBD), and the C-terminal S2 sequence which is relatively conserved between human coronaviruses and SARS-CoV-2 (10).

To better understand the post-vaccination response, we now characterize T-cell, B-cell and IgG responses to S antigenic sequences before and after each dose of mRNA1273 and BNT162b2 vaccines before and over the course of 4 months after starting vaccination in 21 healthy immunocompetent subjects. Single samples from 20 immunocompromised (IC) subjects vaccinated with the mRNA vaccines and Ad26.COV2.S are also evaluated with the same assays.

## METHODS

Subjects were enrolled after informed consent under IRB-approved protocols Pro00045352 and Pro00053511 (Advarra IRB, Columbia, MD) (NCT#04883164). Samples were collected before, two weeks after each vaccine dose, and 4 months after the first vaccine dose in healthy immunocompetent subjects. Samples were collected at any time after completion of vaccination in immunocompromized subjects. All PBL samples were stimulated with overlapping peptide mixtures which represented the less conserved S1 component, the conserved S2 component, and complete S antigenic sequences (JPT Peptides, Germany). The frequencies of CD4+, CD8+ T-cells and CD19+B-cells that expressed CD154 were measured with flow cytometry (6). Monocytic (M-MDSC) and polymorphonuclear (PMN-MDSC) MDSC frequencies were measured as described (6). Anti-RBD and anti-S IgG were measured with ELISA as the optical density at 490 nm (OD_490_). An OD_490_ of 0.45 or greater was considered a positive test for each antibody as described (Xu). Antibody strength was inferred from OD_490_.

## RESULTS

For 21 immunocompetent and 20 IC subjects, mean (SEM) age was 51 (3.7) years and 52 (4.7) years, male: female distribution was 8: 13 and 9: 11, race Caucasian: African-American: Asian was 16: 1: 4 and 18: 1: 1, and vaccine type mRNA1273 (Moderna, Cambridge, MA) vs BNT162b2 (Pfizer, New York, NY) vs Ad26.COV2.S (J and J, NJ) was 11: 9: 0 and 5: 13: 1, respectively. Among immunocompetent subjects, sample collection occurred before vaccination, and at mean±SEM 15±0.2, 47± and 133±4.6, respectively. Single samples from 20 IC subjects were collected earlier, at mean±SEM 101±12.8 days after vaccination, compared with the timing of the last collection from immunocompetent subjects (p=0.029).

### Antibody incidence and strength increases after vaccination followed by early decline in anti-RBD IgG in healthy immunocompetent subjects

The incidence of RBD-IgG and spike-IgG increased from 17/21 and 19/21 two weeks after the first dose, to all 21 subjects being positive for either antibody two weeks after the second dose. By month 4, 1/21 subjects had turned negative for RBD-IgG. This subject has since received a booster. All those with spike-IgG remained positive.

RBD-IgG strength inferred from the mean ± SEM OD_490_ increased significantly from pre-dose to the first (p=2.8E-05) and from the first to the second doses (p=1E-04). This strength decreased significantly between the second dose and month 4 after vaccination, from OD_490_ of 2.87 +/-0.013 to 2.15 +/-0.2, p=0.005. The OD_490_ for anti-S IgG also increased significantly after each dose but was unchanged after the second dose to month 4 (p=0.573, NS). These OD_490_ values were also noteworthy for the larger interindividual variation after the first vaccine dose, and smaller variation after the second dose (Figure 1).

**Figure 1.**
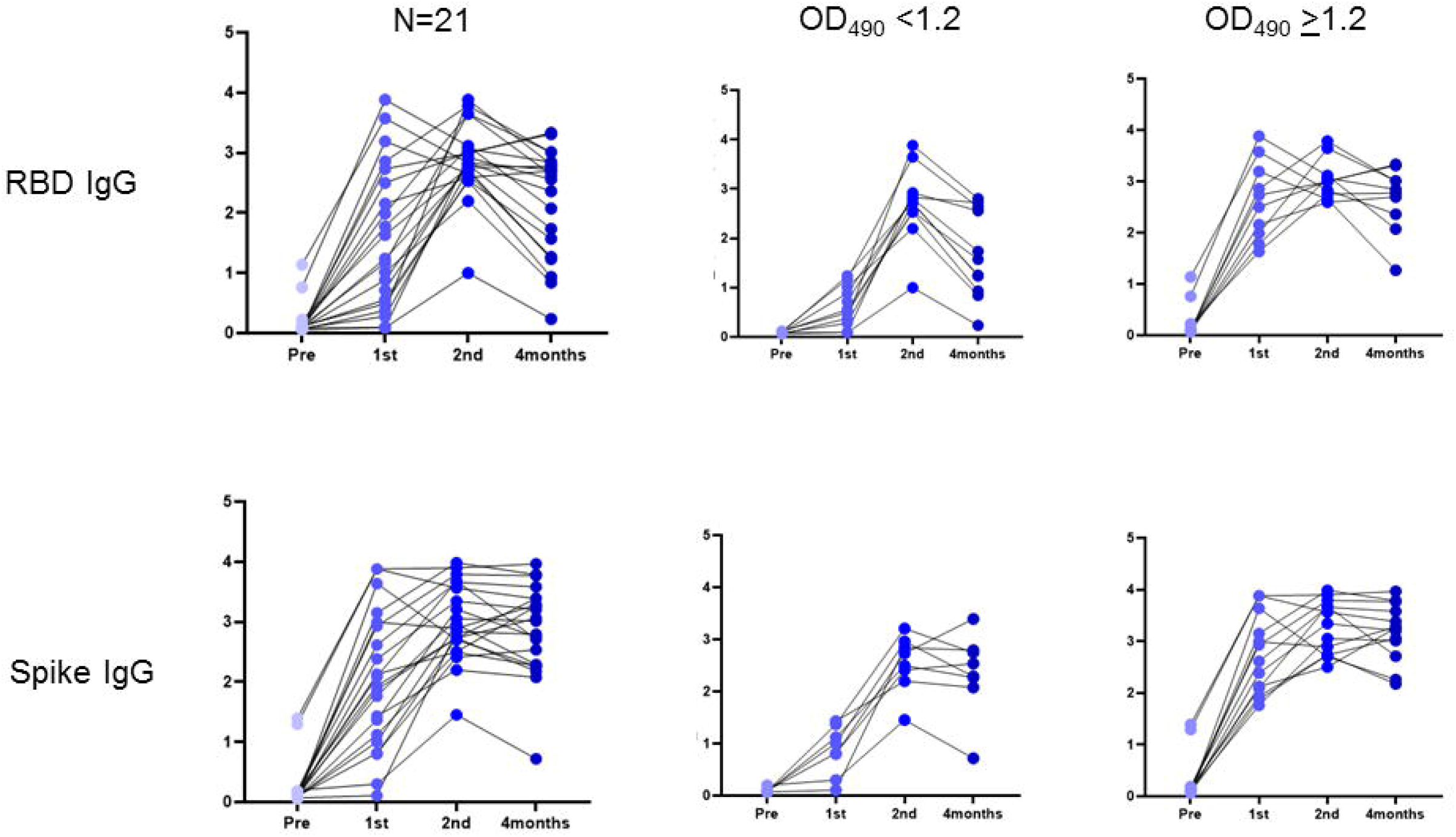
Changes in strength (OD_490_) of anti-RBD IgG (upper panel) and anti-spike IgG (lower panel) for 21 immunocompetent subjects before and after the 1st dose, 2nd dose and 4 months after mRNA vaccination. Also shown are changes for those subjects in each group with OD_490_ <1.2 (middle panels) and >1.2 (right panels).

To evaluate whether the long-term antibody response depended on early antibody kinetics, subjects were arbitrarily divided into those with median OD_490_ less than and greater than the median of 1.2 after the first dose. Four months after vaccination, antibody strength for RBD-IgG (p=0.008) and Spike-IgG (p=0.1, NS) was lower in those subjects with lower median OD_490_ after the first dose, compared with those with higher OD_490_ after the first dose.

### S-reactive T-cells and B-cells co-express IFNγ and IL-6

As reported in a companion study, we first established an assay to examine post-infectious immunity with S-antigen specific T-cells. We established that CD154 is co-expressed with IFNγ, a marker of cytotoxic T-cells, in PBL from 5 healthy human subjects, stimulated overnight with the S peptide mixture (Supplementary Figure 1). Median (range) frequencies of S-reactive T-cells that co-expressed both markers were 3.1% (1.1-10.3), and greatly exceeded S-reactive T-cells that expressed either CD154, 0.2% (0.1-0.2) or IFNγ 0% (0-0.1), respectively. Because nearly all S-reactive IFNγ+T-cells co-express CD154, using the single marker CD154 would overestimate S-reactive IFNγ+T-cells by 0.2% divided by the sum of 0.2% and 3.1% times 100, or 6%. Similarly, in four patients with COVID-19 infection, S-reactive CD154+ IFNγ+T-cells were 0.74% (0.48-0.93), and greatly exceeded CD154+T-cells, 0.03% (0-0.08) or IFNγ+T-cells 0% (0-0) respectively. Because all S-reactive IFNγ+T-cells co-express CD154, using the single marker CD154 in patients with COVID-19 would overestimate S-reactive IFNγ+T-cells by 3.9%. In three healthy human subjects, median (range) frequencies of S-reactive B-cells that co-expressed IL-6 and CD154 were 4.9% (4.7-13.1) and greatly exceeded B-cells that expressed either CD154, 0.4% (0.3-0.4) or IL-6, 0% (0-0), respectively. Because all S-reactive IL-6+B-cells co-express CD154, using the single marker CD154 would overestimate S-antigen reactive IL-6+B-cells by 7.54%. Thus, S-reactive T- and B-cells capture all S-antigen reactive IFNγ+T-cells, and IL-6+B-cells, respectively, and overestimate these cell types by only 3.9-7.5%. In subsequent assays, CD154 was used in lieu of the markers IFNγ an IL-6 in T- and B-cells, respectively.

### B-cells responsive to the conserved S2 increase progressively, while those to SARS-CoV-2 specific S1 spike antigenic sequence increase transiently after vaccination

S2 reactive T- and B-cells are present in many healthy individuals who have not been exposed to SARS-CoV-2 because the S2 component of the spike antigen shares sequence similarities with the human coronaviruses which have caused seasonal flu-like illnesses since the 1960s (10, 11). S2-reactive B-cells decreased after the first vaccine dose compared with pre-vaccination levels but increased progressively at subsequent time points to significantly higher levels (p=0.002, K-W rank sum test) (Figure 2). We have previously categorized the population distribution of S2 reactive B-cell frequencies measured in a sample of 47 independent healthy human subjects who had not been exposed to SARS-CoV-2, into four quartiles, Q1-Q4. Of 21 subjects in the current study, pre-vaccination S2-reactive B-cell frequencies placed 9 subjects in Q1, one subject in Q2, 7 in Q3 and 4 in Q4 (Figure 3). By 4 months after vaccination, S2-reactive B-cell frequencies were in Q3 (n=9) or Q4 (n=11) in 20 of 21 subjects. In the remaining subject, S2-reactive B-cell frequencies were in Q2. None of the 21 vaccinated subjects showed frequencies in the first quartile. S-reactive B-cells also increased steadily from after the first vaccine dose to month 4 (p=0.015). S1-reactive B-cells increased significantly after the second vaccine dose compared with pre-vaccination levels but were undetectable 4 months after vaccination (p=0.019).

**Figure 2.**
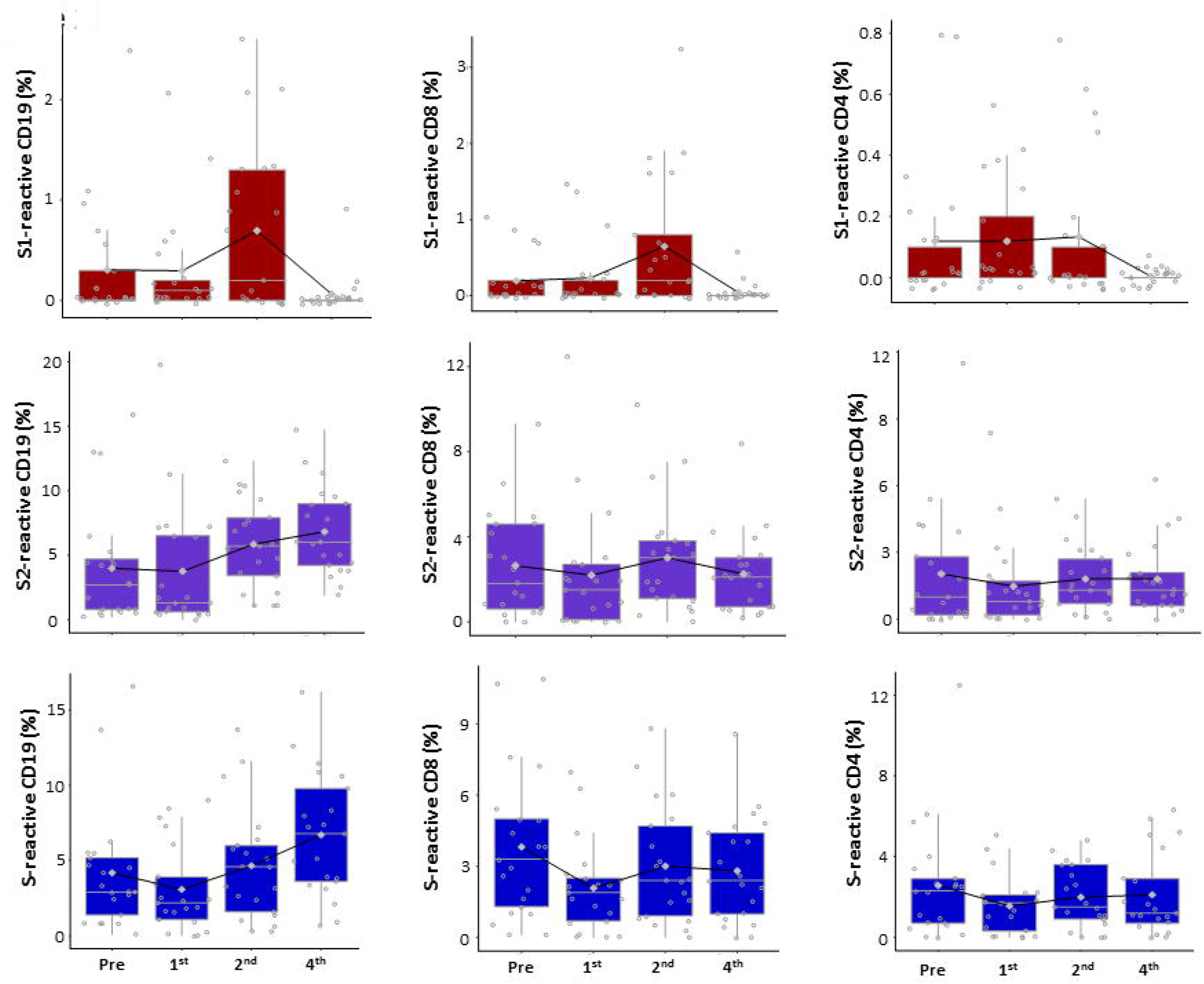
Box plots show S1-reactive (upper row) S2-reactive (middle row) and S-reactive (lower row) CD19+B-cells (left column), CD8+T-cells (middle column) and CD4+T-cells (right column) before (pre) after the first (1st) and 2nd doses and 4 months after starting mRNA vaccination in 21 immunocompetent subjects. Lines connecting boxes for each time point represent mean+/-SEM.

**Figure 3.**
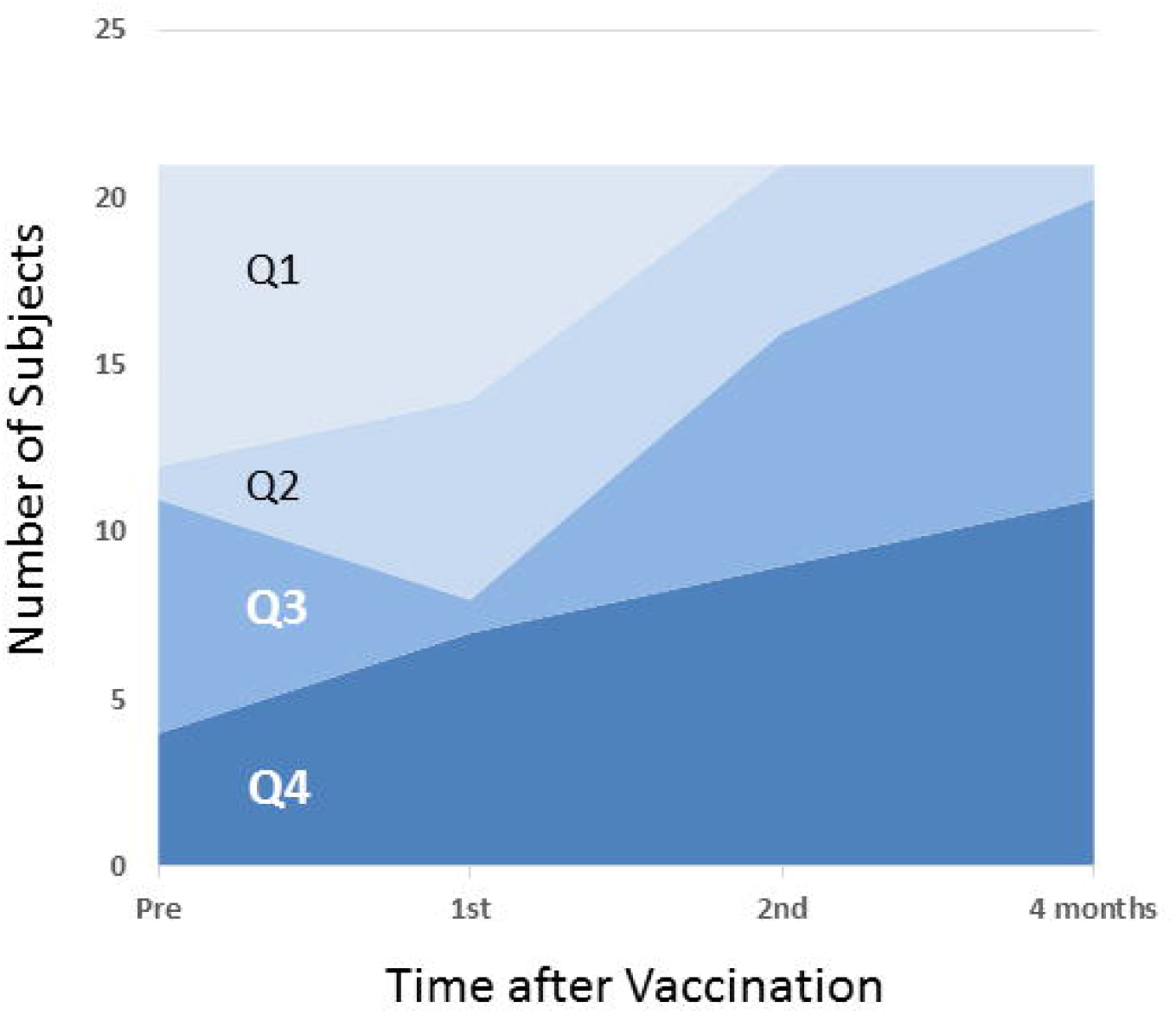
Distribution of quartiles for S2-reactive B-cell frequencies, before and after vaccination in 21 immunocompetent subjects.

### T-cell responsiveness to the SARS-CoV-2-specific S1 sequence but not the conserved S2 sequence increases transiently

CD8 T-cells that were reactive to the SARS-CoV-2-specific spike antigenic sequence S1 were absent before vaccination and increased transiently to levels higher than pre-vaccination levels after the second vaccine dose (Figure 2). S1-reactive cells were not detectable 4 months after vaccination. Differences between observations at all four time points were significant (p=0.002, K-W rank sum test). S2- and S-reactive CD4 and CD8 cells decreased after the first vaccine dose and approached but did not exceed pre-vaccination levels at subsequent timepoints during the first four months (Figure 2).

### Vaccination leads to higher MDSC frequencies

COVID-19 infection is known to induce MDSC which suppress T-cells. Frequencies of monocytic (M-MDSC) and polymorphonuclear (PMN-MDSC) increased significantly after the first vaccine dose and decreased at subsequent time points approaching but not normalizing to pre-vaccination levels (Figure 4). We also found an inverse correlation between M-MDSC frequencies and frequencies of S2-reactive CD19, CD8 and CD4 cells in all 74 samples from all time points in 21 healthy immunocompetent subjects (Figure 4). No such correlations were identified for PMN-MDSC or for S1-reactive or S-reactive CD4, CD8 and CD19 cells.

**Figure 4.**
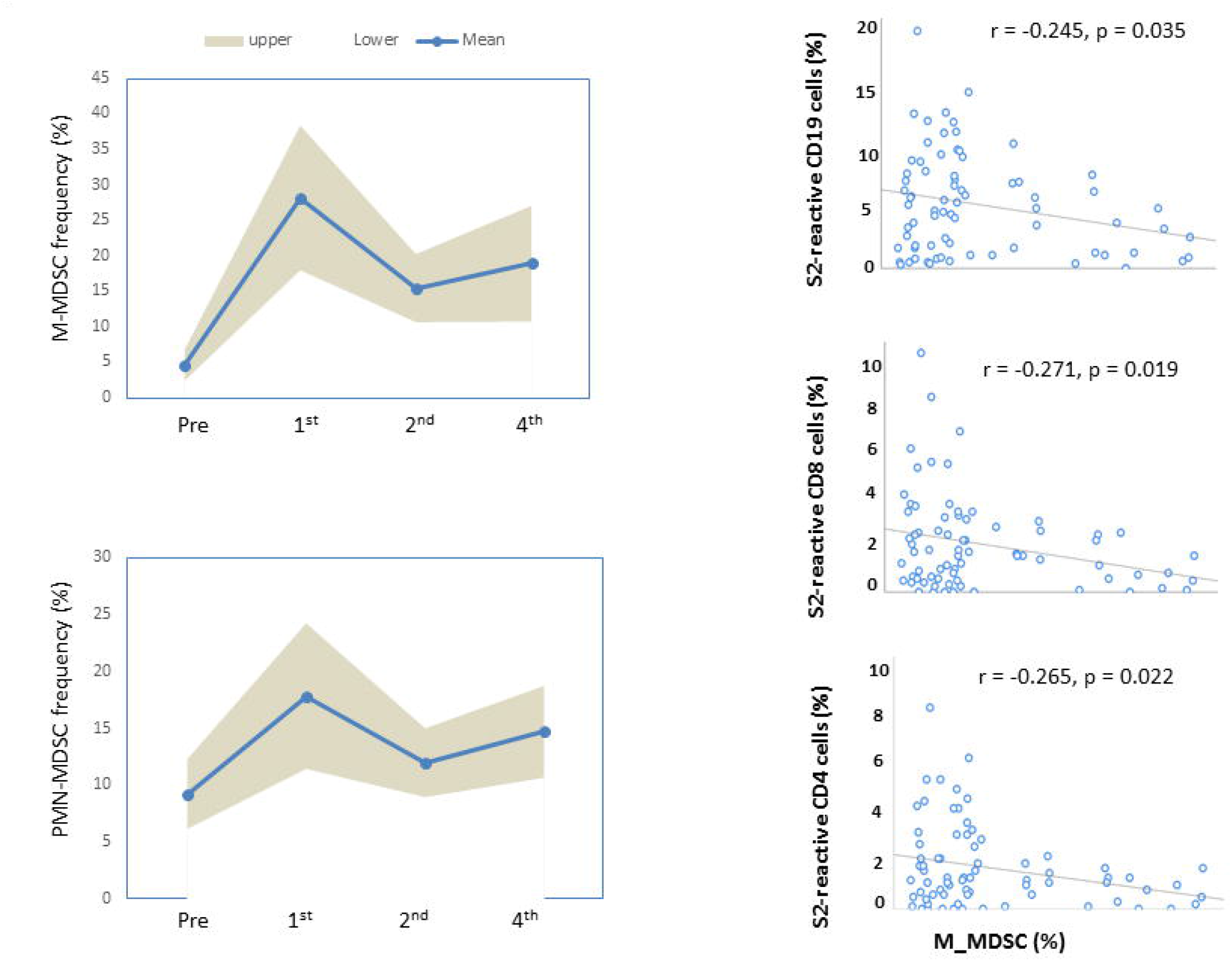
Mean and 95% confidence intervals for serial changes in monocytic M-MDSC (upper left panel) and polymorphonuclear PMN-MDSC (lower left panel). Panels on right show correlations between frequencies of M-MDSC and S2-reactive-CD19, -CD8 and -CD4 cells for all 74 observations before and after vaccination from 21 immunocompetent subjects.

### Immunocompromised subjects have heterogeneous immune responses and enhanced PMN-MDSC

We compared measurements from single samples obtained from 20 IC subjects with those obtained from the final observational timepoint after vaccination in 21 immunocompetent subjects (Figure 5). Antibody-strength reflected by OD_490_ for RBD-IgG and spike-IgG was lower in 20 IC subjects compared with 21 immunocompetent. IC subjects also demonstrated lower frequencies of S-reactive CD4, CD8 and CD19 cells, and elevated frequencies of PMN-MDSC (p<0.05).

**Figure 5.**
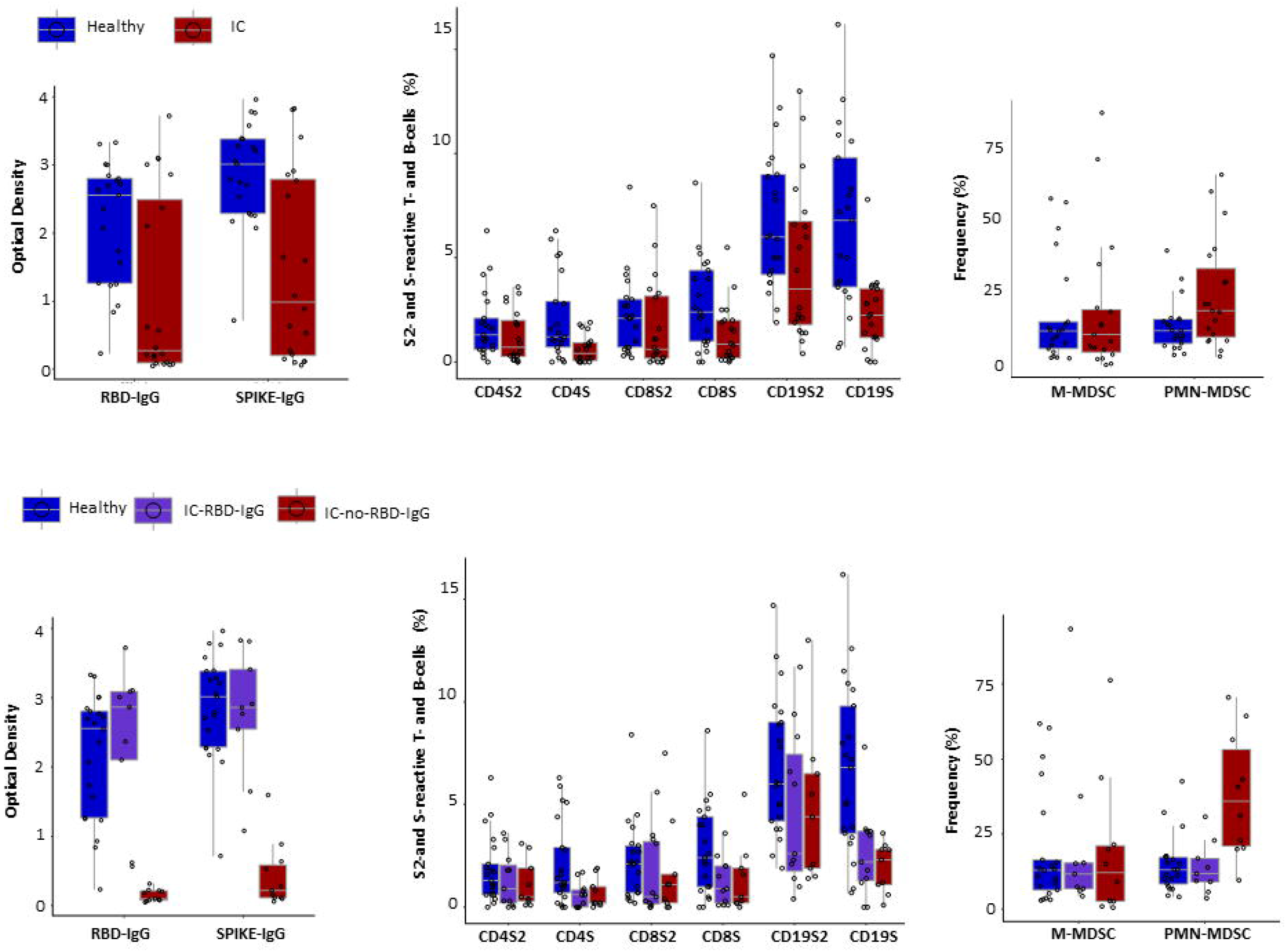
Box plots show strength (OD_490_) of RBD-IgG and Spike-IgG (left column), frequencies of S2-reactive and S-reactive CD4+T-cells, CD8+T-cells and CD19+B-cells (middle column), and frequencies of monocytic and polymorphonuclear M-MDSC and PMN-MDSC (right column. Upper row shows comparisons between 20 healthy immunocompetent subjects with detectable RBD-IgG 4 months after vaccination (blue boxes) and 20 immunocompromised subjects (red boxes, IC) sampled once at 3 or more months after vaccination. In the lower row, immunocompromised subjects have been grouped into those with detectable anti-RBD IgG (purple boxes, IC-RBD-IgG) and those without anti-RBD IgG (red boxes, IC-no RBD-IgG).

To identify the origins of between-group differences in cellular immunity, we compared 20 of 21 immunocompetent subjects with detectable RBD-IgG with 9 IC subjects, also with detectable RBD-IgG and 11 IC subjects without detectable RBD-IgG. S-reactive CD4 and S-reactive B-cell frequencies differed between three groups (p<0.05, Kruskal-Wallis test). In pairwise comparisons, these differences arose from significantly lower frequencies for these two cell types in each IC subgroup, compared with immunocompetent subjects. No such differences were seen between the two IC subgroups. Interestingly, significant differences in PMN-MDSC frequencies between three groups (p=0.002, K-W test) arose from significantly higher PMN-MDSC frequencies in IC patients who failed to make RBD-IgG compared with IC or healthy subjects with RBD-IgG after vaccination (p<0.0004). Differences between three groups in RBD-IgG strength (p=6.9E-06) similarly arose from lower antibody strength in the RBD-IgG negative IC subjects (p<9.3E-07).

## DISCUSSION

Four or more months after vaccination, immunocompetent subjects demonstrate declining RBD-IgG strength and loss of the transient increase in CD8, CD4 and CD19 cell responses to the S1 antigenic sequence, which is more specific for SARS-CoV-2 (Figures 1 and 2). In contrast, spike-IgG strength remains stable between the second dose and the fourth month. The most noteworthy change during this time period is a progressive increase in S2 and S-reactive B-cells (Figures 2 and 3). This change contrasts with a numeric but not statistically significant decrease in S2 and S-reactive CD4 and CD8 T-cells after the first vaccine dose and a return to pre-vaccination levels. After the first vaccine dose, M-MDSC and PMN-MDSC increase (Figure 4). Decreased T-cell responses to the spike protein and increased MDSC frequencies have been observed early after COVID-19 infection (Ashokkumar)(6). Thus, the T-cell response to vaccines recapitulates the effects of COVID-19 infection.

These observations lead to several inferences. *First*, the cellular and antibody response to S1 or its RBD component, which has some sequence-specificity for SARS-CoV-2 is shorter lived compared with Spike-IgG which remains stable until the fourth month. Emerging data at 6 months which will be reported demonstrates stability in the strength of Spike IgG. In this setting, the rationale for early boosters is the declining RBD-IgG response and not the spike-IgG response. *Second*, vaccination augments pre-existing B-cell reactivity to the coronaviruses. Nearly all or 20 of 21 vaccinated immunocompetent subjects achieved S2-reactive B-cell frequencies in the Q3 or Q4 range, 4 months after vaccination compared with 11 of 21 who were in these ranges prior to vaccination (Figure 3).

Sub-phenotyping of S2-reactive B-cells can help to determine whether these cells are memory B-cells, which are known to expand after vaccination, and whether these memory cells augur improved antibody responses to subsequent booster vaccination (Goel)(12). *Third*, the T-cell response to these first vaccinations against the COVID-19 coronavirus appears transient. S1-reactive T-cells declined rapidly, while S2-reactive T-cells were not enhanced above pre-vaccination levels in 4-month follow-up (Figure 2). The T-cell suppressive effect of MDSC which increased significantly after the first vaccination dose provides one explanation (Figure 4). Minimizing the MDSC response to vaccination may increase vaccine efficacy. We have avoided co-stimulation or the use of immunodominant peptides in our cell-based assays, to reduce any likelihood of overinterpreting the T-cell response to SARS-CoV-2 after vaccination of immunocompromised patients (Braun, Grifoni, Sekine, Ashokkumar)(6, 13-15). It is possible that if used in our subject population, these assay modifications may also confirm vaccine-induced reinforcement of pre-existing T-cell immunity to SARS-CoV-2, as has been reported recently (Loyal)(16).

Observations in IC subjects and comparisons between immunocompetent and IC subjects expand on the abovementioned inferences. RBD-IgG was detected in 9 of 20 or 45% while Spike-IgG was detected in 13 of 20 or 65% of vaccinated IC subjects, approximating the incidence of these antibodies in previous reports from vaccinated IC populations (Figure 5). The absence of RBD-IgG in 11 of 20 IC subjects was associated with enhanced circulating PMN-MDSC frequencies in this sub cohort compared with immunocompetent subjects (p<0.0004). These associations are supported by studies in which PMN-MDSC inhibit B-cell proliferation and antibody production (Lelis, Knier)(17, 18). However, MDSC are also known to expand in IC populations such as transplant recipients, and cancer patients (Lee, Feng, Cassetta)(19-21). This expansion was not apparent in vaccinated IC subjects with RBD-IgG, in whom M-MDSC and PMN-MDSC frequencies approximated those seen in immunocompetent subjects. Notably, over half or 11 of 21 immunocompetent subjects received mRNA1273 vaccination compared with a fourth or 5 of 20 IC subjects (p=0.11, NS). Also, 11 IC subjects without RBD-IgG included six solid organ transplant recipients on Tacrolimus, steroids, or mycophenolate mofetil (Supplementary Table 1), compared with one such recipient in IC with RBD-IgG. In the RBD-IgG-negative IC group, belatacept which inhibits donor-specific antibody development in transplant recipients and velcade which is used to treat antibody mediated rejection in transplant patients were used in a subject with multiple myeloma and a kidney transplant recipient, respectively. To better understand the post-vaccine antibody response in IC subjects, the role of MDSC and immunosuppressive regimens will need ongoing reassessment.

Vaccinated IC subjects also revealed a heterogenous cellular response to the conserved S2 and the complete S antigenic sequences suggesting varying immunogenicity of the spike antigenic sequences (Figure 5). The frequencies of S2-reactive CD4, CD8 and CD19 B-cells were not significantly different between immunocompetent subjects and the two IC sub cohorts, one of which failed to make RBD-IgG. This observation is of interest because S2-reactive B-cell frequencies rose progressively until the fourth month after vaccination. Thus, similar ranges of S2-reactive B-cell frequencies in immunocompetent and IC subjects at an average of 132 and 101 days after vaccination, respectively, would imply a post vaccination augmentation in both types of subjects, and also suggest that pre-existing immunity due to previous exposure to human coronaviruses may offer some protection in both types of subjects. Serial observations are needed in IC subjects. In contrast, S-reactive CD4+T-cell and B-cell frequencies were significantly lower in the IC sub cohorts compared with immunocompetent subjects but similar between the two IC sub cohorts, one of which failed to produce RBD-IgG after vaccination. The dynamic range of S-reactive cell frequencies is larger than S2-reactive cells and may better discern differences between groups. Other reasons may include a greater immunogenicity of the S1 and S2 components together, than either component alone. In support of this inference, T- and B-cells responsive to stimulation with the isolated S1 antigenic mixture were not detected 4 months after vaccination, suggesting that this component may be less immunogenic by itself. This heterogeneity suggests that T- and B-cells respond variably to the spike antigenic components. Further, cellular and humoral immunity does not go hand in hand in IC subjects. Thus, protective immunity may be an aggregate of many known and unknown responses to vaccination.

In conclusion, COVID-19 vaccination induces a transient increase in cellular and antibody responses to the SARS-CoV-2-specific S1 spike antigenic sequence. A sustained antibody response to the complete spike protein that is stable for at least 4 months is accompanied by a progressive increase in B-cells that are responsive to this spike protein, and its component that is conserved between the human coronaviruses and SARS-CoV-2. Although half of the IC subjects in this study failed to seroconvert after vaccination, T-cell and B-cell reactive to the conserved S2 component of the spike protein may provide cellular immunity at levels similar to those seen in IC and immunocompetent subjects who seroconverted after vaccination.

## Data Availability

All data that underlie the results reported in this Article (including study protocol) on individual participants will be made available to researchers who provide a methodologically sound proposal to the corresponding author.

## Conflict of Interest

Some assays described here are included in University of Pittsburgh Patent 9606019, author: RS, which is licensed exclusively to Plexision, in which the University and RS own equity. CA is a paid consultant to Plexision. The remaining authors declare that the research was conducted in the absence of any commercial or financial relationships that could be construed as a potential conflict of interest.

## Author Contributions

Conceptualization: RS, CA, Methodology: RS, CA, Assays: CA, BS, MS, Data accrual, review, quality assessment and organization: BS, MS, ES, Statistical analysis and graphics: VS, Patient recruitment: BN, AM, PS, AR, MS. Writing: Original draft: RS. Writing: Final draft, review and editing: RS, CA, VS, AM, BS, MS, ES, SS, BN.

## Funding

NSF 2033307 for assay standardization and reference ranges.

## Acknowledgments

Ms Kathleen Smith and Carole Ann Capezzuto for manuscript preparation.

## Data Availability Statement

All data needed to evaluate the conclusions in the paper are present in the paper and/or the Supplementary materials. Additional data and code related to this paper can be requested from the authors

## Figure Caption

**Supplementary Figure 1.** From Ashokkumar, 2021, *bioRxiv* (2021) doi:10.1101/2021.05.03.442371. Flow cytometry scatterplots show a) expresson of CD154 and IFNγ in S-reactive and PMA-reactive T-cells and b) expression of CD154 and IL-6 in S-reactive and PMA-reactive B-cells.

## Notes

### Clinical Trial

NCT#04883164

### Funding Statement

NSF 2033307 for assay standardization and reference ranges. Plexision for internal funding

### Author Declarations

Subjects were enrolled after informed consent under IRB-approved protocols Pro00045352 and Pro00053511 (Advarra IRB, Columbia, MD) (NCT#04883164).

## REFERENCES

1. Bates, T. A., Leier, H. C., Lyski, Z. L., Goodman, J. R., Curlin, M. E., Messer, W. B., et al.: Age-Dependent Neutralization of SARS-CoV-2 and P.1 Variant by Vaccine Immune Serum Samples. JAMA, 326(9), 868–9 (2021) doi:10.1001/jama.2021.11656

2. Schwarz, T., Tober-Lau, P., Hillus, D., Helbig, E. T., Lippert, L. J., Thibeault, C., et al.: Delayed Antibody and T-Cell Response to BNT162b2 Vaccination in the Elderly, Germany. Emerg Infect Dis, 27(8), 2174–2178 (2021) doi:10.3201/eid2708.211145

3. Boyarsky, B. J., Werbel, W. A., Avery, R. K., Tobian, A. A. R., Massie, A. B., Segev, D. L., et al.: Antibody Response to 2-Dose SARS-CoV-2 mRNA Vaccine Series in Solid Organ Transplant Recipients. JAMA, 325(21), 2204–2206 (2021) doi:10.1001/jama.2021.7489

4. Kennedy, N. A., Lin, S., Goodhand, J. R., Chanchlani, N., Hamilton, B., Bewshea, C., et al.: Infliximab is associated with attenuated immunogenicity to BNT162b2 and ChAdOx1 nCoV-19 SARS-CoV-2 vaccines in patients with IBD. Gut, 70(10), 1884–1893 (2021) doi:10.1136/gutjnl-2021-324789

5. Agha, M., Blake, M., Chilleo, C., Wells, A. and Haidar, G.: Suboptimal response to COVID-19 mRNA vaccines in hematologic malignancies patients. medRxiv (2021) doi:10.1101/2021.04.06.21254949

6. Ashokkumar, C., Rohan, V., Kroemer, A. H., Rao, S., Mazariegos, G., Higgs, B. W., et al.: Impaired T-cell and antibody immunity after COVID-19 infection in chronically immunosuppressed transplant recipients. bioRxiv (2021) doi:10.1101/2021.05.03.442371

7. Agrati, C., Sacchi, A., Bordoni, V., Cimini, E., Notari, S., Grassi, G., et al.: Expansion of myeloid-derived suppressor cells in patients with severe coronavirus disease (COVID-19). Cell Death Differ, 27(11), 3196–3207 (2020) doi:10.1038/s41418-020-0572-6

8. Schulte-Schrepping, J., Reusch, N., Paclik, D., Baßler, K., Schlickeiser, S., Zhang, B., et al.: Severe COVID-19 Is Marked by a Dysregulated Myeloid Cell Compartment. Cell, 182(6), 1419–1440.e23 (2020) doi:10.1016/j.cell.2020.08.001

9. O’Connor, M. A., Rastad, J. L. and Green, W. R.: The Role of Myeloid-Derived Suppressor Cells in Viral Infection. Viral Immunol, 30(2), 82–97 (2017) doi:10.1089/vim.2016.0125

10. Jaimes, J. A., André, N. M., Chappie, J. S., Millet, J. K. and Whittaker, G. R.: Phylogenetic Analysis and Structural Modeling of SARS-CoV-2 Spike Protein Reveals an Evolutionary Distinct and Proteolytically Sensitive Activation Loop. J Mol Biol, 432(10), 3309–3325 (2020) doi:10.1016/j.jmb.2020.04.009

11. Kahn, J. S. and McIntosh, K.: History and recent advances in coronavirus discovery. Pediatr Infect Dis J, 24(11 Suppl), S223–7, discussion S226 (2005) doi:10.1097/01.inf.0000188166.17324.60

12. Goel, R. R., Apostolidis, S. A., Painter, M. M., Mathew, D., Pattekar, A., Kuthuru, O., et al.: Distinct antibody and memory B cell responses in SARS-CoV-2 naïve and recovered individuals following mRNA vaccination. Sci Immunol, 6(58) (2021) doi:10.1126/sciimmunol.abi6950

13. Braun, J., Loyal, L., Frentsch, M., Wendisch, D., Georg, P., Kurth, F., et al.: SARS-CoV-2-reactive T cells in healthy donors and patients with COVID-19. Nature, 587(7833), 270–274 (2020) doi:10.1038/s41586-020-2598-9

14. Grifoni, A., Weiskopf, D., Ramirez, S. I., Mateus, J., Dan, J. M., Moderbacher, C. R., et al.: Targets of T Cell Responses to SARS-CoV-2 Coronavirus in Humans with COVID-19 Disease and Unexposed Individuals. Cell, 181(7), 1489–1501.e15 (2020) doi:10.1016/j.cell.2020.05.015

15. Sekine, T., Perez-Potti, A., Rivera-Ballesteros, O., Strålin, K., Gorin, J. B., Olsson, A., et al.: Robust T Cell Immunity in Convalescent Individuals with Asymptomatic or Mild COVID-19. Cell, 183(1), 158–168.e14 (2020) doi:10.1016/j.cell.2020.08.017

16. Loyal, L., Braun, J., Henze, L., Kruse, B., Dingeldey, M., Reimer, U., et al.: Cross-reactive CD4(+) T cells enhance SARS-CoV-2 immune responses upon infection and vaccination. Science (2021) doi:10.1126/science.abh1823

17. Lelis, F. J. N., Jaufmann, J., Singh, A., Fromm, K., Teschner, A. C., Pöschel, S., et al.: Myeloid-derived suppressor cells modulate B-cell responses. Immunol Lett, 188, 108–115 (2017) doi:10.1016/j.imlet.2017.07.003

18. Knier, B., Hiltensperger, M., Sie, C., Aly, L., Lepennetier, G., Engleitner, T., et al.: Myeloid-derived suppressor cells control B cell accumulation in the central nervous system during autoimmunity. Nat Immunol, 19(12), 1341–1351 (2018) doi:10.1038/s41590-018-0237-5

19. Lee, Y. S., Zhang, T., Saxena, V., Li, L., Piao, W., Bromberg, J. S., et al.: Myeloid-derived suppressor cells expand after transplantation and their augmentation increases graft survival. Am J Transplant, 20(9), 2343–2355 (2020) doi:10.1111/ajt.15879

20. Feng, S., Cheng, X., Zhang, L., Lu, X., Chaudhary, S., Teng, R., et al.: Myeloid-derived suppressor cells inhibit T cell activation through nitrating LCK in mouse cancers. Proc Natl Acad Sci U S A, 115(40), 10094–10099 (2018) doi:10.1073/pnas.1800695115

21. Cassetta, L., Bruderek, K., Skrzeczynska-Moncznik, J., Osiecka, O., Hu, X., Rundgren, I. M., et al.: Differential expansion of circulating human MDSC subsets in patients with cancer, infection and inflammation. J Immunother Cancer, 8(2) (2020) doi:10.1136/jitc-2020-001223

