## Supplementary figures and images for "Cellular and Antibody Immunity after COVID-19 Vaccination at >4-Month Follow Up in Immunocompetent and Immunocompromised Subjects"

### Sup Figure 1

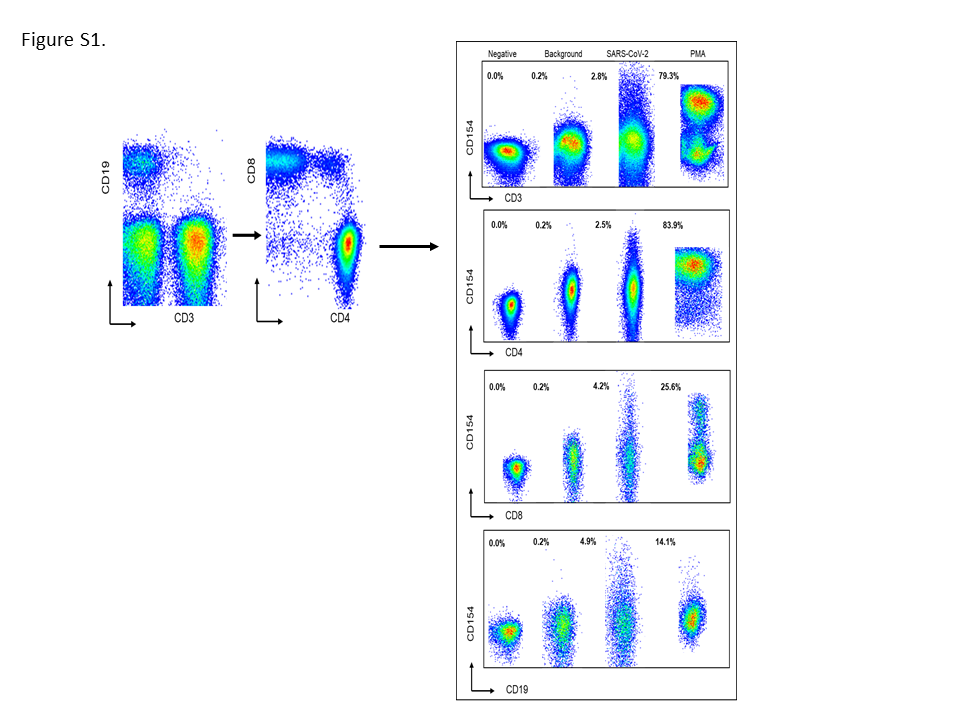
