## Supplementary material for "Cellular and Antibody Immunity after COVID-19 Vaccination at >4-Month Follow Up in Immunocompetent and Immunocompromised Subjects": Sup_Table 1

**Supplementary Table 1.**

**Characteristics of 20 immunocompromised sampled cross-sectionally after COVID-19 vaccination.**

| **Condition** | **Medication** | **Vaccine** | **RBD-IgG** | **SPIKE-IgG** |
| --- | --- | --- | --- | --- |
| **Multiple myeloma** | Velcade | mRNA1273 | Negative | Negative |
| **Kidney Transplant** | Mycophenolate mofetil, Belatacept, Prednisone. | mRNA1273 | Negative | Negative |
| **Kidney Transplant** | Tacrolimus, Belatacept | BNT162b | Negative | Negative |
| **Heart transplant** | Tacrolimus, steroid | BNT162b | Negative | Negative |
| **Combined variable immune deficiency** | Human Immune globulin (Cuvitru) | BNT162b | Negative | Negative |
| **Intestine transplant** | Tacrolimus, steroids | BNT162b | Negative | Negative |
| **Liver transplant** | Rapamune, Prednisone | BNT162b | Negative | Negative |
| **Rheumatoid Arthritis** | Leflunomide, Medroxyprogesterone acetate | Ad26.COV2.S | Negative | Positive |
| **Inflammatory bowel disease** | Mycophenolate mofetil, Simponi-Aria (TNF inhibitor), steroids, leflunomide | BNT162b | Negative | Positive |
| **Rheumatoid Arthritis** | Humira | BNT162b | Negative | Positive |
| **Kidney Transplant** | Tacrolimus, mycophenolate mofetil | BNT162b | Negative | Positive |
| **Gynecological cancer** | off chemotherapy | mRNA1273 | Positive | Positive |
| **Psoriatic Arthritis** | Ixekizumab (anti-IL-17 antibody) | mRNA1273 | Positive | Positive |
| **Psoriatic Arthritis** | Certolizumab (anti-TNF), Methotrexate, prednisone | mRNA1273 | Positive | Positive |
| **Bone marrow transplant** | off immunosuppression | BNT162b | Positive | Positive |
| **Multiple Sclerosis** | Dimethyl Fumarate | BNT162b | Positive | Positive |
| **Liver transplant** | Tacrolimus | BNT162b | Positive | Positive |
| **Prostate cancer** | off chemotherapy | BNT162b | Positive | Positive |
| **Chronic myelogenous leukemia** | Dasatinib | BNT162b | Positive | Positive |
| **Rheumatoid Arthritis** | Methotrexate, Remicade | BNT162b | Positive | Positive |
